# Prevalence and Predictors of Late presentation to HIV/AIDS care in Uvira, South Kivu Province, Democratic Republic of the Congo

**DOI:** 10.1101/2025.10.14.25338035

**Authors:** Kyambikwa Bisangamo Célestin, Zawadi Lubala Jeanne, Asima Katumbi Florentin, Nessrin Ahmed El-Nimr

## Abstract

**Background:** Late presentation (LP) to HIV/AIDS care remains a major barrier to effective treatment outcomes and the reduction of HIV transmission, particularly in resource-limited settings. In the Democratic Republic of the Congo (DRC), despite national initiatives to expand HIV testing and antiretroviral therapy (ART) coverage, many people living with HIV (PLHIV) still enter care at advanced stages of disease progression. This study aimed to determine the prevalence and identify the predictors of LP to HIV/AIDS care in Uvira.

**Methods:** A cross-sectional study was conducted from August to October 2024 among PLHIV attending ART clinics in Uvira. A total of 275 participants were proportionally recruited from all 10 ART sites using consecutive sampling. Data were collected through structured interviews and clinical record reviews. LP was defined as WHO clinical stage III/IV or a CD4 count below 350 cells/mm³ at entry into care. Data analysis was performed using Epi Info and SPSS software. Bivariate and multivariate logistic regression analyses were conducted to identify independent predictors of LP, with significance set at p < 0.05.

**Results:** The prevalence of LP was 51.3%. Independent predictors of LP included alcohol intake (AOR = 2.01; 95% CI: 1.09–3.70), age above 24 years (AOR = 3.17; 95% CI: 1.13–8.93), monthly income ≥200 USD (AOR = 2.02; 95% CI: 1.06–3.85), no formal education (AOR = 3.09; 95% CI: 1.28–7.48), fear of disclosure (AOR = 5.23; 95% CI: 2.03–13.5), and non-use of condoms (AOR = 2.38; 95% CI: 1.07–5.31).

**Conclusion:** LP to HIV care remains highly prevalent in Uvira, highlighting persistent challenges in early diagnosis and linkage to care. Strengthening community awareness, expanding HIV testing, and addressing socioeconomic and behavioral barriers are crucial to enhancing early engagement in care and improving treatment outcomes across the DRC.

## Background

Acquired Immune Deficiency Syndrome (AIDS) is an infectious disease caused by Human Immunodeficiency Virus (HIV). Despite efforts for its prevention and control, AIDS continues to be a major global health problem [1]. By of the end of 2023, 39.9 million people worldwide were living with HIV [2]. Africa remains the most affected region, with nearly 1 in every 30 adults (3.4%) living with HIV and accounting for more than two-thirds of the people living with HIV (PLHIV) worldwide [3]. HIV prevalence in the Democratic Republic of Congo (DRC) is estimated to be 0.7% amongst adults aged 15–49 years. Approximately 470,000 people in DRC are living with HIV, with 91% of them receiving antiretroviral therapy (ART) [4].

Since 2016, the World Health Organization (WHO) has recommended that all newly diagnosed patients with HIV should start antiretroviral therapy (ART) on the day of diagnosis. [5]. ART, when started on time, has been a remarkable achievement in recent medicine, significantly reducing morbidity and mortality among individuals infected with HIV and significantly reducing the risk of HIV transmission [6,7].

Globally, late presentation (LP) for HIV/AIDS care is recognized as a public health problem. It is estimated that between 25% and 35% of PLHIV worldwide present late for HIV/AIDS care [8]. In the DRC, particularly in Kinshasa, LP prevalence has decreased from 70.7% in 2013 to 46.5% in 2017 and 23.4% in 2020 [9]. LP was defined as persons who presented for care with a CD4+ T cell count of less than 350 cells/mm^3^ [10]. According to the WHO, LP is defined as patients presenting to care with HIV clinical stage III or IV [11]. LP to HIV/AIDS care is associated with a number of short- and long-term consequences, including poor treatment outcomes, increased healthcare costs, ART drug resistance, early development of opportunistic infections, and early mortality [12–14]. On the other hand, PLHIV who arrive late for care are also more likely to spread the virus to healthy people because their serostatus is unknown [15]. Similarly, PLHIV who arrive late for care incur significant financial costs compared to those who arrive earlier [16–18].

Late presentation for HIV care remains a major challenge in achieving optimal treatment outcomes and epidemic control. Several studies have identified multiple predictors influencing delayed diagnosis and linkage to care. Older age and male sex are consistently associated with late presentation, as older individuals often undergo less frequent testing and men show lower health-seeking behaviour and engagement with health services [19,20]. Socioeconomic deprivation, including poverty, unemployment, and low educational attainment, further limits access to HIV testing and treatment [15]. Stigma, discrimination, and insufficient social support remain powerful deterrents to early care-seeking [21]. In addition, low perceived risk of infection, limited testing coverage, and missed clinical opportunities contribute to diagnostic delays [22,23]. Migrant status, rural residence, and poor geographic access to health facilities are also linked to late presentation [21,24]. Variations across key populations and treatment interruptions among previously diagnosed individuals exacerbate the problem [15]. Addressing these determinants requires tailored interventions that expand HIV testing, reduce stigma, and strengthen decentralized and community-based care systems. To improve health outcomes, guide healthcare professionals’ treatment plans, and inform policymakers about emerging health issues, identifying factors that predict LP is essential [8].

Although several studies have been carried out on the various forms of LP to HIV/AIDS care in developing countries and few parts of the DRC, there is limited information on predictors of LP to HIV/AIDS care in South Kivu Province, particularly in the Uvira health zone. This study is one of the first to comprehensively assess LP to HIV care in Uvira, South Kivu, using a broad range of sociodemographic, behavioral, and clinical factors and multivariate analysis to identify independent predictors. Findings provide valuable local data to guide targeted interventions. Therefore, the present study aimed to estimate the prevalence and to identify possible predictors of LP to HIV care in Uvira Health Zone, South Kivu Province, DRC.

## Methods

### Study design and settings

A cross-sectional study was conducted among PLHIV attending ART clinics in the Uvira health zone from August to October 2024.

The Uvira health zone is a crossroads, facilitating transactions with several countries, especially Burundi, Rwanda, Tanzania, and Zambia. It has a population of 411,438, an area of 1,306 km^2^ and a population density of 344 inhabitants per km^2^. The Uvira health zone is one of 34 health zones in the province of South Kivu. It is located in the territory and city of Uvira, in the south of the province of South Kivu. It is bordered to the North by the Ruzizi rural health zone via the Kawizi river, to the South by the Nundu rural health zone via the Kambekulu river, to the East by the Republic of Burundi via Lake Tanganyika, and to the West by the Hauts-Plateaux rural health zone via the Mitumba mountain range. The Uvira health zone has 10 health facilities with integrated HIV/AIDS services. These are Kavinvira Health Center, Uvira General Referral Hospital, Kasenga/CEPAC General Hospital, Kalundu Catholic Health Center, Kalundu / CEPAC Health Center, Kigongo Health Center, Nyamianda Hospital Center, Kabindula Health Center, SOS Kala Health Center, and Rombe Health Center [25].

### Study participants and sampling

The study was conducted among patients with HIV newly diagnosed by clinicians using standard diagnostic protocol of WHO [5]. Inclusion criteria were adult patients with HIV (aged 18 years and above) who were registered and followed up at health facilities for at least one year after entry into care, with at least two clinical visits, and who had recorded relevant initial diagnostic information (WHO clinical stage at entry into care) [9].

All 10 ART clinics in the Uvira health zone were included in the study. The sample size was calculated using Epi Info version 7.2.3.1. Based on a prevalence of LP of 23.4% [9], and 5% acceptable margin of error, the minimum required sample size at 95% confidence level was calculated to be 275 patients with HIV. The sample was proportionately allocated to the 10 ART clinics according to the total number of patients registered in each clinic. Patients with HIV were consecutively recruited until the completion of the required sample size.

### Data collection

Data were collected in face-to-face interviews conducted by trained interviewers. A predesigned structured interviewing questionnaire was used to collect data in three main sections: Sociodemographic characteristics (age, gender, marital status, level of education, employment status, monthly income, and living arrangements), lifestyle factors (mode of acquiring HIV, attending TH or prayer room, knowledge of HIV attendance, travel time to ART clinic, alcohol intake, smoking, number of sexual partners, and condom use), perceptions and clinical history related to HIV infection (perception that HIV is curable and preventable, fear of disclosure and stigma, HIV testing initiative, symptoms at first HIV diagnosis, chronic illness, and pre-testing counseling).

For each patient, details of the WHO clinical stage at the time of diagnosis were recorded. Participants with LP were defined as people with HIV/AIDS confirmed at WHO stage 3 or 4 at the time of diagnosis or CD4 cell count < 350 cells/mm^3^. Participants without LP were defined as people with HIV/AIDS confirmed at WHO stage 1 or 2 at the time of diagnosis or CD4 cell count ≥ 350 cells/mm^3^ [8,9].

### Data management and statistical analysis

Data were checked for completeness during the data collection process. Data were entered into KoboToolbox, cleaned, and coded in Microsoft Excel 2016. The collected data were analyzed using Epi Info version 7.2.3.1 and IBM SPSS (Statistical Package for the Social Sciences) version 25.

Categorical variables were summarized using frequencies and percentages. Median and interquartile range were used to summarize quantitative variables based on the distribution of the data. Bivariate comparisons of categorical variables were assessed using Pearson chi-square test and crude odds ratio. When Pearson chi-square test was invalid, Fisher exact test was used for 2x2 tables. A multivariate logistic regression model was used to calculate the adjusted odds ratio and identify significant predictors of LP to HIV care as the dependent variable. The significance level was set at a p-value of less than 0.05.

### Ethical considerations

This study was conducted in strict accordance with international ethical guidelines for research involving human participants. Ethical approval was obtained from the Ethics Committee of the Bukavu Higher Institute of Medical Techniques, Bukavu, South Kivu, DRC (IRB N°: ISTM-BKV/CRPS/CIES/MLM/011/2024). Written informed consent was obtained from each participant prior to data collection. Participation was entirely voluntary, and individuals were informed of their right to withdraw from the study at any time without any consequences. Anonymity and confidentiality of all participants were rigorously maintained throughout the research process.

## Results

The median age was 35 years (IQR: 15.5), with the majority (70.5%) aged between 25 and 50 years. Most participants were female (71.3%) and married (46.2%), while 30.9% were single. Regarding education, over half (50.5%) had attained secondary or high school education, and 14.2% had tertiary-level qualifications. A substantial proportion (71.3%) were employed, though nearly half (46.9%) reported a monthly income below 100 USD. In terms of living arrangements, 46.9% lived with their spouse, 28.7% lived alone, and 24.4% lived with family members. (Supplementary table 1)

The majority (90.5%) reported acquiring HIV through heterosexual transmission. Only 12.7% reported attending traditional healers or prayer rooms, and most participants (91.6%) were aware of HIV care attendance requirements. Among those receiving ART, 55.2% traveled more than one hour to reach the clinic. Regarding lifestyle behaviors, 38.2% reported alcohol consumption, whereas cigarette or shisha smoking was rare (2.6%). Slightly more than half (54.2%) had more than one sexual partner, and among them, 63.8% never used condoms, while 36.2% reported occasional use. (Supplementary table 2)

A large majority (96.7%) believed that HIV is not curable, while 66.9% perceived it as preventable. Fear of disclosure was reported by 85.1% of participants, and an even higher proportion (95.6%) expressed fear of stigma. Most participants (68.7%) reported that their HIV testing was initiated by health workers. At the time of diagnosis, 63.6% presented with symptoms, and only a small minority (4.7%) reported having another chronic illness. Notably, only 18.2% received pre-testing counseling, indicating limited access to comprehensive pre-diagnosis support. (Supplementary table 3) **Prevalence of LP to HIV/AIDS care**

The prevalence of LP among PLHIV in Uvira health zone was 51.3% (Figure 1).

**Figure 1.**
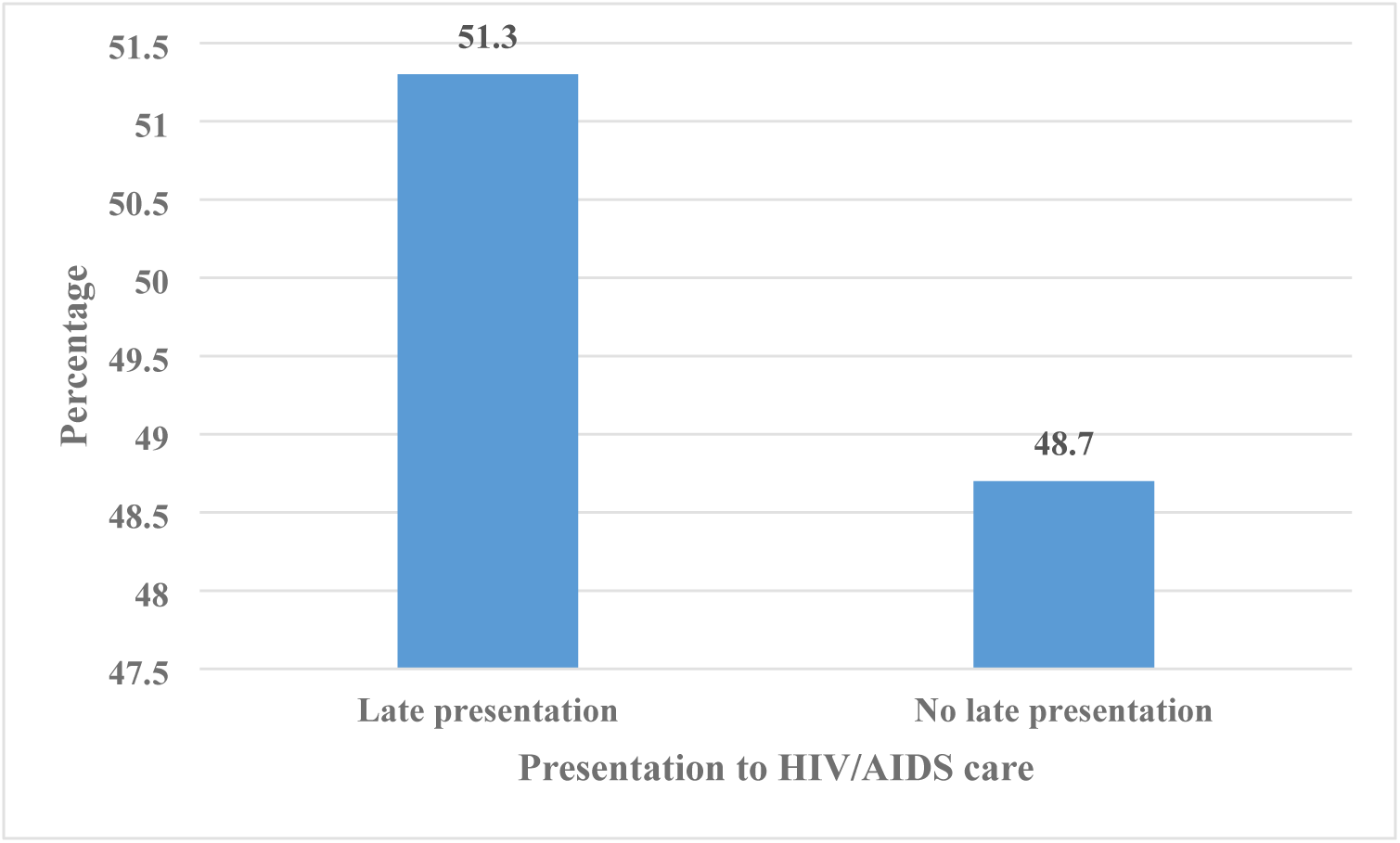
Prevalence of LP to HIV/AIDS care among study participants, Uvira, 2024 Distribution of participants according to their personal and demographic characteristics and their presentation to HIV/AIDS care, Uvira, 2024.

Table 1 presents the associations between sociodemographic characteristics and LP to HIV/AIDS care. Age was significantly associated with LP: participants aged 25–50 years and those older than 50 years were more likely to present late compared to those aged 18–24 years. Educational level was also significantly related to LP, with higher odds among participants with no formal education, primary, and secondary education, compared to those with tertiary education. Additionally, participants earning more than 200 USD per month were nearly twice as likely to present late, and those living alone had significantly higher odds of LP.

**Table 1.** Distribution of participants according to their personal and demographic characteristics and their presentation to HIV/AIDS care, Uvira, 2024.

| Variable | Presentation to HIV/AIDS care |  | OR (95% CI) | *p-value* |
| --- | --- | --- | --- | --- |
|  | LP (%) | No LP (%) |  |  |
| Age Group (Years) |  |  |  |  |
| 18-24 | 15 (31.3) | 33 (68.7) | Ref |  |
| 25-50 | 101 (52.0) | 93 (48.0) | 2.39 (1.22-4.68) | 0.009* |
| > 50 | 25 (75.8) | 8 (24.2) | 6.87 (2.52-18.7) | <0.001* |
| Gender |  |  |  |  |
| Male | 37 (46.8) | 42 (53.2) | Ref |  |
| Female | 104 (53.1) | 92 (46.9) | 1.28 (0.76-2.16) | 0.349 |
| **Marital Status** |  |  |  |  |
| Currently single | 81 (44.7) | 67 (55.3) | 1.35 (0.84-2.17) | 0.216 |
| Currently married | 60 (47.2) | 67 (52.8) | Ref |  |
| **Level of Education** |  |  |  |  |
| No formal | 17 (73.9) | 6 (26.1) | 6.37 (2.01-20.2) | <0.001* |
| Primary School | 39 (52.7) | 35 (47.3) | 2.51 (1.10-5.69) | 0.026* |
| Secondary School | 73 (52.5) | 66 (47.5) | 2.49 (1.17-5.31) | 0.016* |
| Tertiary School | 12 (30.8) | 27 (69.2) | Ref |  |
| **Employment Status** |  |  |  |  |
| Employed | 106 (54.1) | 90 (45.9) | 1.48 (0.87-2.50) | 0.142 |
| Not employed | 35 (44.3) | 44 (55.7) | Ref |  |
| **Monthly Income (US \$)** | | | | |
| < 100 | 61 (47.3) | 68 (52.7) | Ref |  |
| 100-200 | 22 (41.5) | 31 (58.5) | 0.79 (0.41-1.51) | 0.477 |
| > 200 | 58 (62.4) | 35 (37.6) | 1.85 (1.07-3.18) | 0.026* |
| **Living Arrangements** |  |  |  |  |
| Living alone | 51 (64.6) | 28 (35.4) | 2.54 (1.30-4.95) | 0.006* |
| Husband/wife | 62 (48.1) | 67 (51.9) | 1.29 (0.71-2.34) | 0.403 |
| Families | 28 (41.8) | 39 (58.2) | Ref |  |

OR, odds ratio; CI, confidence interval; \* significant ( $p < 0.05$ ); IQR, interquartile range.

### Distribution of lifestyle factors among PLHIV according to their LP for HIV/AIDS care, Uvira, 2024

The distribution of lifestyle factors among PLHIV according to their LP for HIV/AIDS care is shown in Table 2. Participants who reported alcohol consumption were significantly more likely to present late compared to non-drinkers. Having more than one sexual partner was also strongly associated with LP. Among participants with multiple sexual partners, inconsistent condom use further increased the likelihood of LP, with those who never used condoms showing nearly threefold higher odds. Other lifestyle factors did not show significant associations with LP for HIV/AIDS care.

**Table 2.**
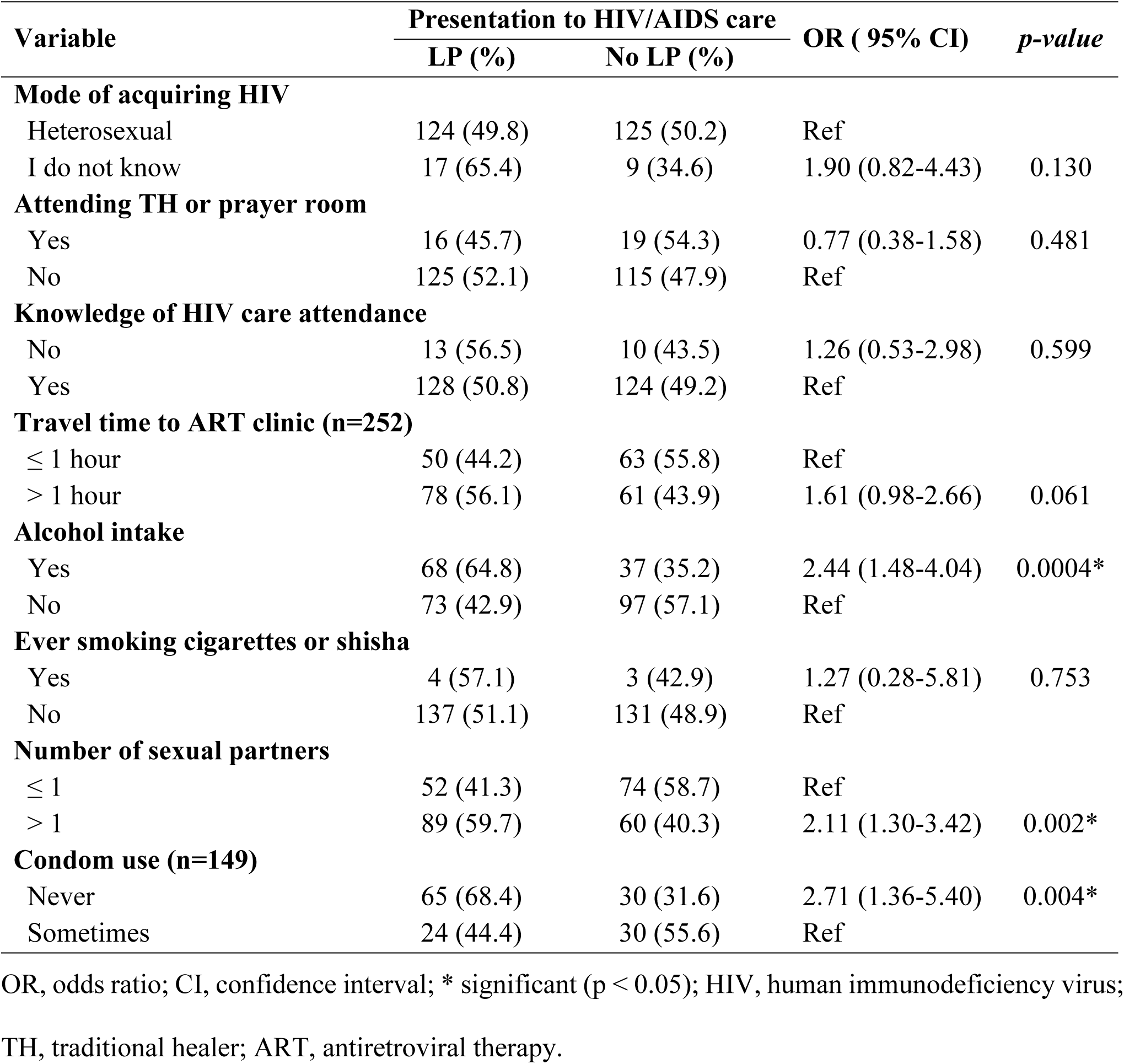
Distribution of lifestyle factors among PLHIV according to their LP for HIV/AIDS care, Uvira, 2024.

### Distribution of perceptions and clinical history related to HIV infection associated with LP to HIV/AIDS care

Table 3 summarizes the relationship between perceptions and clinical history related to HIV infection and LP to HIV/AIDS care. Participants reporting fear of disclosure were significantly more likely to present late to care. Similarly, having symptoms at the time of HIV diagnosis and the presence of a chronic illness were also significantly associated with LP. Conversely, perceptions that HIV is curable or preventable, fear of stigma, HIV testing initiative, and receipt of pre-testing counseling did not show statistically significant associations (p > 0.05).

**Table 3.** Distribution of perceptions and clinical history related to HIV infection associated with LP.

| Variable | Presentation to HIV/AIDS care |  | OR ( 95% CI) | *p-value* |
| --- | --- | --- | --- | --- |
|  | LP (%) | No LP (%) |  |  |
| **Perception that HIV is curable** |  |  |  |  |
| No | 138 (51.9) | 128 (48.1) | 2.16 (0.53-8.80) | 0.225 |
| Yes | 3 (33.3) | 6 (66.7) | Ref |  |
| **Perception that HIV is preventable** |  |  |  |  |
| No | 49 (53.8) | 42 (46.2) | 1.17 (0.70-1.93) | 0.548 |
| Yes | 92 (50.0) | 92 (50.0) | Ref |  |
| **Fear of disclosure** |  |  |  |  |
| Yes | 134 (57.3) | 100 (42.7) | 6.51 (2.77-15.3) | <0.0001* |
| No | 7 (17.1) | 34 (82.9) | Ref |  |
| **Fear of stigma** |  |  |  |  |
| Yes | 133 (50.6) | 130 (49.4) | 0.51 (0.15-1.74) | 0.379 |
| No | 8 (66.7) | 4 (33.3) | Ref |  |
| **HIV testing initiative** |  |  |  |  |
| Health workers | 98 (51.8) | 91 (48.2) | 1.08 (0.65-1.79) | 0.776 |
| Myself/friend | 43 (50.0) | 43 (50.0) | Ref |  |
| **Symptoms at first HIV diagnosis** |  |  |  |  |
| Yes | 98 (56.0) | 77 (44.0) | 1.69 (1.03-2.77) | 0.038* |
| No | 43 (43.0) | 57 (57.0) | Ref |  |
| **Chronic illness** |  |  |  |  |
| Yes | 11 (84.6) | 2 (15.4) | 5.58 (1.21-25.7) | 0.020* |
| No | 130 (49.6) | 132 (50.4) | Ref |  |
| **Pre-testing counseling** |  |  |  |  |
| No | 113 (50.2) | 112 (49.8) | 0.79 (0.43-1.47) | 0.459 |
| Yes | 28 (56.0) | 22 (44.0) | Ref |  |

OR, odds ratio; CI, confidence interval; HIV, human immunodeficiency virus; \* significant ( $p < 0.05$ ).

### Multivariate logistic regression analysis of predictors of late presentation (LP) to HIV/AIDS care

The multivariate logistic regression analysis presented in Table 4 identifies the independent predictors of LP to HIV/AIDS care. After adjustment for potential confounders, fear of disclosure emerged as the strongest predictor (AOR = 5.23; 95% CI: 2.03–13.5; p = 0.0006), indicating a markedly higher likelihood of delayed care among those fearing disclosure. Other significant predictors included alcohol intake (AOR = 2.01; p = 0.025), age > 24 years (AOR = 3.17; p = 0.029), monthly income ≥ 200 USD (AOR = 2.02; p = 0.032), lack of formal education (AOR = 3.09; p = 0.012), and non-use of condoms (AOR = 2.38; p = 0.033). Other variables, including marital status, living arrangement, chronic illness, number of sexual partners, and presence of symptoms at diagnosis, were not significantly associated with LP (p > 0.05).

**Table 4.** Multivariate logistic regression analysis of predictors of LP to HIV/AIDS care, Uvira, 2024.

| Predictors of late presentation | AOR | (95% CI) | p-value |
| --- | --- | --- | --- |
| Alcohol intake (Yes) | <u>2.011</u> | <u>(1.09-3.70)</u> | <u>0.0248*</u> |
| Age group (> 24 years old) | <u>3.170</u> | <u>(1.13-8.93)</u> | <u>0.0289*</u> |
| Monthly income (≥ 200 USD) | <u>2.022</u> | <u>(1.06-3.85)</u> | <u>0.0319*</u> |
| Living arrangement (Living alone) | 1.692 | (0.87-3.30) | 0.1224 |
| Marital status (Divorced) | 1.322 | (0.53-3.30) | 0.5487 |
| Chronic illness (Yes) | 4.391 | (0.77-25.1) | 0.0959 |
| Level of education (No formal) | <u>3.094</u> | <u>(1.28-7.48)</u> | <u>0.0121*</u> |
| Number of sexual partners (> 1) | 0.907 | (0.42-1.95) | 0.8030 |
| Fear of disclosure (Yes) | <u>5.234</u> | <u>(2.03-13.5)</u> | <u>0.0006**</u> |
| Condoms use (No) | <u>2.383</u> | <u>(1.07-5.31)</u> | <u>0.0334*</u> |
| Symptoms at first HIV diagnosis (Yes) | 1.411 | (0.77-2.60) | 0.2687 |

AOR, adjusted odds ratio; CI, confidence interval; \*significant (p < 0.05)

## Discussion

Early initiation of ART plays a pivotal role in improving the health outcomes of PLHIV and in reducing onward transmission, particularly among individuals who are unaware of their serostatus. However, LP to HIV/AIDS care remains a major public health concern, especially in resource-limited settings where early diagnosis and timely initiation of ART are critical for achieving optimal outcomes. Assessing the prevalence and identifying the predictors of LP are essential steps toward developing targeted interventions that promote earlier engagement in HIV/AIDS care. In the context of Uvira, understanding these determinants is particularly important for informing context-specific strategies aimed at reducing HIV-related morbidity, mortality, and transmission.

The prevalence of LP in Uvira was 51.3%, which is lower than the rates reported in Cameroon (89.7%, 2018) [10], Zambia (76.7%, 2024) [8], and South Africa (79.0%, 2017) [26], but comparable to those observed in Kosovo (50.6%, 2023) [23], Ethiopia (52.9%, 2019) [27], Brazil (52.7%, 2024) [28], and Kenya (53.0%, 2019) [29]. However, it exceeds the estimates documented in Kinshasa, DRC (23.4%, 2021) [9], Belgium (38.3%, 2024) [15], Uganda (29.6%, 2019) [30], Ghana (28.6%, 2024) [19], Tanzania (42.4%, 2024) [31], and Mozambique (47.0%, 2022) [32]. These disparities may be attributed to contextual differences such as socioeconomic status, cultural factors, HIV prevalence, testing strategies, and accessibility of health services [33].

The present study found that age, education, and income were significantly associated with late presentation (LP) to HIV care. Consistent with previous reports, older participants (24–50 years and >50 years) were more likely to present late than those aged ≤24 years [9,19,33–35]. This may reflect the misconception that older adults are at lower risk of HIV infection, leading to reduced awareness and less frequent testing.

Educational level was also associated with LP: Patients with LP had 3 times the likelihood of not having any formal education compared to patients who had no LP (AOR=3.09; 95% CI: 1.28–7.48; p=0.0121). This finding was in line with earlier studies [31,36,37]. Limited health literacy, misinformation, stigma, and barriers to navigating health services may underlie this association.

Unexpectedly, LP was significantly related to higher monthly income (≥200 USD). Similar findings have been reported in Ethiopia and other sub-Saharan contexts, where employment or higher socioeconomic status has been associated with delayed care entry [38]. Possible explanations include low risk perception, competing professional obligations, and reluctance to seek services due to stigma. Late presentation to HIV/AIDS care was also significantly associated with alcohol use and non-use of condoms (p<0.05). Similar findings have been reported in Ethiopia, Kenya, and the Democratic Republic of the Congo. A pooled analysis of Ethiopian studies identified frequent alcohol use as a strong predictor of LP [27], while research in Kenya showed that heavy drinking was linked to unawareness of HIV serostatus, lack of ART use, and viremia, all markers of delayed engagement in care [39]. Although evidence directly connecting condom non-use to LP is limited, its frequent occurrence among new HIV cases suggests a possible role in delayed testing and care-seeking, particularly when compounded by alcohol use, stigma, or denial.

The current results showed that fear of disclosure, symptoms at first diagnosis, and presence of chronic illness were significantly associated with LP to HIV/AIDS care. These findings are consistent with earlier research. Fear of disclosing HIV status has been widely documented as a barrier to timely linkage and retention in care. Studies from sub-Saharan Africa report that individuals who anticipate negative reactions from family or community often delay seeking HIV services due to concerns about rejection or violence [40,41]. Similarly, chronic illness and symptomatic presentation at diagnosis typically reflect a delayed entry into care, when patients only seek medical attention after experiencing advanced HIV-related symptoms or comorbidities [42,43]. This pattern highlights gaps in early detection and proactive health-seeking behaviors.

Conversely, perceptions that HIV is curable or preventable, fear of stigma, HIV testing initiative, and pre-testing counseling were not significantly associated with LP. Although stigma is often reported as a critical determinant of delayed HIV care [44,45], the absence of significance here may indicate contextual differences: stigma may manifest more subtly, or individuals might separate general fears of societal stigma from the personal act of disclosure. Similarly, perceptions about HIV cure or prevention may influence general attitudes toward HIV but not necessarily the timing of seeking care. Furthermore, HIV testing initiative (whether voluntary or provider-initiated) and pre-test counseling did not affect late presentation, possibly reflecting improvements in routine HIV testing policies and standardized counseling that reduce disparities in care linkage [46].

Overall, the results suggest that interpersonal and health status–related factors (disclosure, symptoms, chronic illness) exert stronger influence on late presentation than knowledge-based or procedural factors (perceptions, testing mode, counseling). Interventions should therefore prioritize stigma reduction at the level of disclosure, community education about asymptomatic HIV, and screening for comorbidities to promote earlier entry into care.

In multivariate analysis, alcohol intake, age (>24 years), higher income (>200 USD/month), lack of formal education, fear of disclosure, and non-use of condoms were found to be independent predictors of LP to HIV/AIDS care.

The association between alcohol use and LP may be explained by behavioral and psychosocial mechanisms. Alcohol consumption has been linked to lower health-seeking behavior, reduced adherence to care, and increased risky practices, which can delay HIV testing and timely linkage to treatment [41,42].

Late presentation to HIV care was more frequent among participants aged over 24 years compared to younger ones. This is consistent with findings from several sub-Saharan studies, where younger individuals, particularly those enrolled through antenatal care or provider-initiated testing, are diagnosed earlier, while older adults often delay until symptomatic [43].

Interestingly, higher monthly income was also associated with LP. Although financial stability is usually protective, in some contexts, individuals with higher income may perceive themselves as healthier, rely on private care, or face stigma in workplace settings, contributing to delays in seeking HIV services [45].

The role of education was clear. Limited health literacy has been consistently associated with poor HIV knowledge, reduced risk perception, and lower uptake of testing and care services [40,46].

The strongest predictor identified was fear of disclosure, which increased the odds of LP more than fivefold. This finding aligns with prior studies that highlight disclosure-related stigma as one of the most significant barriers to timely care and treatment [40,44,45]. Addressing disclosure fears through community support and stigma reduction interventions remains critical.

Finally, non-use of condoms was associated with LP. This may reflect broader risk-taking behaviors, lower HIV awareness, and reduced engagement with prevention and testing programs among individuals who do not regularly use condoms [41].

The limitations of this study include its cross-sectional design, reliance on self-reported data, and the single-site setting, which may limit the generalizability of the findings. In addition, the study did not account for potentially influential factors such as mental health status, distance to healthcare facilities, and broader health system determinants.

## Conclusion and recommendations

Late presentation to HIV care remains a major public health challenge in Uvira, where its prevalence remains high. LP was independently associated with older age, lack of formal education, higher income, alcohol use, non-use of condoms, and fear of disclosure. Among these, fear of disclosure was the strongest predictor, underscoring the central role of stigma in delaying care. These findings highlight that LP is shaped not only by clinical and demographic factors but also by behavioral and psychosocial determinants. Addressing LP is therefore critical to improving treatment outcomes and reducing HIV transmission in this setting.

Recommendations include strengthening community education on asymptomatic HIV, early testing, and timely ART initiation for adults over 24 years; implementing stigma-reduction and disclosure-support programs; integrating alcohol reduction and risk-reduction counseling; expanding adult-focused testing beyond antenatal settings; promoting health literacy among those with no formal education; and routinely screening for comorbidities to enable earlier HIV diagnosis and care linkage.

## Data Availability

No data sets were generated or analyzed in the current study.

## Abbreviations

LP: Late Presentation
HIV: Human Immunodeficiency Virus
PLHIV: People Living with HIV
AIDS: Acquired Immune Deficiency Syndrome

## Acknowledgements

We thank all study participants

## Author contributions

All authors contributed substantially to the study, including its conception, design, data collection, analysis, and interpretation. They participated in drafting, revising, or critically reviewing the manuscript, approved the final version for publication, agreed on the target journal, and accept responsibility for all aspects of the work.

## Data availability

No data sets were generated or analyzed in the current study.

## Declarations

### Ethics approval and consent to participate

This study was conducted in accordance with the Declaration of Helsinki and received approval from the Ethics Committee of the Bukavu High Institute of Medical Techniques (ISTM-Bukavu), the Central Office of the Uvira Health Zone, and the health facilities providing care for people living with HIV. All procedures adhered to international research ethics guidelines.

### Consent for publication

Not applicable.

## Supplementary Tables

**Table 1.** Sociodemographic characteristics of the study participants, Uvira, 2024.

| Variable | n=275 | % |
| --- | --- | --- |
| Age Group (Years) |  |  |
| 18-24 | 48 | 17.5 |
| 25-50 | 194 | 70.5 |
| > 50 | 33 | 12 |
| Median (IQR) | 35.0 (15.5) |  |
| Gender |  |  |
| Male | 79 | 28.7 |
| Female | 196 | 71.3 |
| Marital Status |  |  |
| Single | 85 | 30.9 |
| Married | 127 | 46.2 |
| Divorced | 37 | 13.5 |
| Widowed | 26 | 9.4 |
| Level of Education |  |  |
| No formal | 23 | 8.4 |
| Primary School | 74 | 26.9 |
| Secondary / High School | 139 | 50.5 |
| Tertiary School | 39 | 14.2 |
| Employment Status |  |  |
| Employed | 196 | 71.3 |
| Not employed | 79 | 28.7 |
| Monthly Income (US \$) | | |
| < 100 | 129 | 46.9 |
| 100-200 | 53 | 19.3 |
| > 200 | 93 | 33.8 |
| Living Arrangements |  |  |
| Living alone | 79 | 28.7 |
| Husband/wife | 129 | 46.9 |
| Families | 67 | 24.4 |

**Table 2.** Lifestyle factors of people living with HIV, Uvira, 2024.

| Variable | n=275 | % |
| --- | --- | --- |
| **Mode of acquiring HIV** |  |  |
| Heterosexual | 249 | 90.5 |
| I do not know | 26 | 9.5 |
| **Attending TH or prayer room** |  |  |
| Yes | 35 | 12.7 |
| No | 240 | 87.3 |
| **Knowledge of HIV care attendance** |  |  |
| No | 23 | 8.4 |
| Yes | 252 | 91.6 |
| **Travel time to ART clinic (n=252)** |  |  |
| ≤ 1 hour | 113 | 44.8 |
| > 1 hour | 139 | 55.2 |
| **Alcohol intake** |  |  |
| Yes | 105 | 38.2 |
| No | 170 | 61.8 |
| **Ever smoking cigarettes or shisha** |  |  |
| Yes | 7 | 2.6 |
| No | 268 | 97.4 |
| **Number of sexual partners** |  |  |
| ≤ 1 | 126 | 45.8 |
| > 1 | 149 | 54.2 |
| **Condom use (n=149)** |  |  |
| Never | 95 | 63.8 |
| Sometimes | 54 | 36.2 |

**Table 3.** Perceptions and clinical history related to HIV infection among PLHIV, Uvira, 2024.

| Variable | n=275 | % |
| --- | --- | --- |
| **Perception that HIV is curable** |  |  |
| No | 266 | 96.7 |
| Yes | 9 | 3.3 |
| **Perception that HIV is preventable** |  |  |
| No | 91 | 91 (33.1) |
| Yes | 184 | 66.9 |
| **Fear of disclosure** |  |  |
| Yes | 234 | 85.1 |
| No | 41 | 14.9 |
| **Fear of stigma** |  |  |
| Yes | 263 | 95.6 |
| No | 12 | 4.4 |
| **HIV testing initiative** |  |  |
| Health workers | 189 | 68.7 |
| Myself/friend | 86 | 31.3 |
| **Symptoms at first HIV diagnosis** |  |  |
| Yes | 175 | 63.6 |
| No | 100 | 36.4 |
| **Chronic illness** |  |  |
| Yes | 13 | 4.7 |
| No | 262 | 95.3 |
| **Pre-testing counseling** |  |  |
| No | 225 | 81.8 |
| Yes | 50 | 18.2 |

